# Longitudinal Weekly Surveillance of Respiratory Viruses and the Nasal Microbiome in Children Under Five (MINNE-LOVE Study)

**DOI:** 10.64898/2026.03.05.26347454

**Authors:** Olivia Toles, Benjamin Jorgenson, Ayan Sheikdon, Sabrina Arif, Vicky Yang, Alexander Johnson, Rick Jansen, Bart Theelen, Beth K. Thielen

**Affiliations:** Department of Pediatrics, University of Minnesota Medical School, Minneapolis, MN, United States of America; University of Minnesota School of Public Health, Division of Epidemiology and Community Health, University of Minnesota, Minneapolis, MN, USA; University of Chicago; Bioinformatics and Computational Biology graduate program, University of Minnesota, Minneapolis, MN, USA; Biostatistics Core, Masonic Cancer Center, University of Minnesota, Minneapolis, MN, USA

**Keywords:** Target-enriched high-throughput nucleotide sequencing, 16S rRNA-based microbial profiling, longitudinal cohort study, respiratory tract microbiota, pediatric respiratory tract infections

## Abstract

Early childhood is a critical period for both nasal microbiome development and susceptibility to respiratory viral infections. While prior cross-sectional and limited longitudinal studies suggest that the nasal microbiome is shaped by both host and environmental factors, high-frequency longitudinal data linking microbial dynamics and respiratory viral disease in young children remain sparse.

**Methods:** We conducted the MINNE-LOVE prospective longitudinal cohort study of children under 5 years of age with weekly symptom surveillance and parent-collected anterior nasal swabs. Total nucleic acid was extracted and analyzed for nasal microbiome composition by 16S rRNA sequencing on short- and long-read platforms and viral pathogen detection by target-enriched metagenomic sequencing. Microbiome diversity, community structure, viral detection patterns, and correlations among dominant bacterial taxa were assessed over time.

**Results:** Within individuals, microbiome composition was relatively stable over time, with acute shifts observed. We detected a broad array of respiratory viruses, including frequent viral co-detections and prolonged detection of select viruses across multiple weeks.

**Conclusion:** In this longitudinal cohort of young children undergoing high-frequency sampling, we demonstrated the feasibility of multiomic assessment of nasal microbial communities. Key bacterial ecological relationships described in prior cross-sectional studies were recapitulated using dense temporal sampling. Target-enriched sequencing enhanced the range of viral pathogen detection, including co-infections and prolonged viral shedding. Full-length 16S long-read sequencing enabled clinically relevant species-level resolution not achievable with short-read approaches. These findings highlight the value of intensive longitudinal cohort designs for defining host–virus–microbiome interactions in early childhood and informing future mechanistic and interventional studies.

**Key points:** We conducted weekly symptom assessment, nasal microbiome profiling, and respiratory virus detection by target-enrichment metagenomics in four Minnesota preschool children. We demonstrated high symptom and viral burden, intraindividual microbiome stability, and improved taxonomic resolution with long-read 16S gene sequencing.

## Introduction

Lower respiratory tract infections (LRTIs) remain a major cause of global morbidity and mortality despite substantial reductions in bacterial disease achieved through vaccination and antimicrobial therapy [1]. With the widespread adoption of molecular diagnostics, viral pathogens are now recognized as major contributors to this burden [2,3], yet preventive and therapeutic options for most respiratory viruses remain limited or poorly durable [4]. Addressing the persistent burden of RTIs requires a deeper understanding of the host and microbial factors driving disease pathogenesis.

A central challenge in respiratory viral disease is the striking heterogeneity in clinical outcomes, even among individuals infected with the same pathogen, with variability in the tendency to progress from limited upper respiratory tract disease to more dangerous lower respiratory tract disease [5]. If the biological drivers underlying interindividual variability in viral respiratory tract infection severity were better understood, this knowledge could enable identification of actionable targets to modulate disease course and improve clinical outcomes. Traditional experimental and clinical models provide only partial insight into the determinants of interindividual variability in respiratory viral disease severity. Commonly used murine systems lack the genetic diversity [6,7], immunologic history [8], and burden of co-infections that shape human outcomes [9], while hospital-based human studies are inherently biased toward severe presentations and often fail to capture pre-illness and early-life determinants. Community-based longitudinal human studies address key limitations capturing the real-world heterogeneity in host genetics, prior immunity and co-infection and by enabling direct observation of pre-infection states, acute viral infection events, and recovery trajectories that are not discernible in cross-sectional designs.

Among factors amenable to study in longitudinal studies is the respiratory microbiome. Several lines of evidence implicate the upper respiratory microbiome as a potentially modifiable contributor to this variability [10,11]. First, cross-sectional studies have identified distinct microbiome profiles that are associated with the presence of respiratory tract disease [12–15]. Second, longitudinal birth cohorts demonstrate that early-life microbiome composition predicts future respiratory health, with stable communities enriched in *Corynebacterium* sp. or *Dolosigranulum pigrum* associated with fewer infections and profiles dominated by *Haemophilus* spp. or *Streptococcus* spp. linked to increased risk [14,16–18]. Third, recent studies suggest that respiratory infections with both viral pathogens and bacterial pathobiont species are more strongly associated with disease symptoms than either alone [19]. Fourth, mechanistic studies further show that commensal airway bacteria can modulate antiviral immunity through type I and III interferon pathways, shaping early viral control, inflammatory responses, and vaccine immunogenicity. Commensal bacteria enhance tonic interferon signaling [20], protect against severe influenza in animal models [21,22], and influence influenza vaccine immunogenicity in humans [23]. Taken together, these findings position the respiratory microbiome as a potential determinant of viral disease heterogeneity. While previous studies have characterized the development of the pediatric upper respiratory microbiome [24–26] and its associations with specific pathogens [27,28], only one longitudinal study has examined correlations between the respiratory microbiome and viral infections in infants under one year of age [15], and no studies to date have extended this approach to older children.

Building on these microbiome–virus associations, recent advances in molecular diagnostics now enable a broader assessment of the respiratory virome, extending beyond the limited scope of conventional multiplex PCR panels [15,29] through the use of next-generation sequencing allow integrated, high-resolution profiling of both viral pathogens and the surrounding microbial community. Hybrid-capture, target-enrichment platforms support simultaneous analysis of bacterial and viral taxa and improve characterization of co-infections [30]. This includes both a broad range of respiratory and systemic viruses in children, including not only common acute pathogens captured by standard clinical multiplex panels but also chronic or intermittently shed viruses such as cytomegalovirus (CMV), Epstein Barr virus (EBV), (human herpes virus 6 (HHV-6), and parvovirus B19 that are typically not assessed in routine diagnostic testing. Leveraging these tools, we conducted a longitudinal study of children under 5 years that captured baseline data on factors known to influence microbiome (mode of delivery, early life nutrition, household composition) as well as weekly nasal swabs and symptom surveillance across a typical winter respiratory season. To specifically test the utility of novel sequencing-based detection platforms, we piloted paired 16S rRNA sequencing with target-enriched viral genome sequencing. This multi-omics approach provides an opportunity to define patterns of microbiome stability and perturbation in early childhood, identify microbial predictors of viral infection and disease severity, and generate mechanistic hypotheses to guide the development of microbiome-informed preventive and therapeutic strategies.

## Methods

### Study Site and Recruitment

The Microbiome Influences on Nursery Nasal Environment–Longitudinal Observations of Viral Epidemiology (MINNE-LOVE) Study is a longitudinal, community-based pediatric cohort study conducted through the University of Minnesota (UMN), a major academic medical and research center in Minneapolis, and its surrounding communities. Participants under five years of age were recruited through multiple pathways, including referrals from UMN-affiliated pediatric clinics and community outreach through study flyers and social media posts, a REDCap [31] pre-screening website, and the Driven to Discover (D2D) program at the Minnesota State Fair and county fairs. All specimens and questionnaire data were collected remotely with informed written e-consent from a parent or guardian [32], under a protocol approved by the University of Minnesota Institutional Review Board (STUDY00014148). Only a single parent’s consent was required due to minimal risk; assent was not obtained because all participants were under five years of age. Participants were eligible if they were <60 months old at the start of the respiratory viral season (designated November 1 of year of participation) and had a parent or guardian able to provide consent and collect weekly nasal swabs. Individuals were excluded if the investigator judged any condition to pose safety concerns or compromise data quality, or if a contraindication to routine mucosal specimen collection was present. To enhance engagement and retention, families were provided with a participant-friendly website and individualized viral detection and microbiome reports.

### Clinical data

Following consent, a parent/guardian completed a baseline questionnaire on household demographics and medical history relevant to microbiome composition [33,34]. Weekly follow-up questionnaires were administered electronically via REDCap to the parent’s primary email address. Weekly digital questionnaires collected information on household environment (number of children and adults, daycare or school attendance, pets, smoking in the home, type of residence) and participant symptoms, including fever, cough, nasal congestion, wheeze, or gastrointestinal illness. Questionnaire also captured missed daycare/school, antibiotic use, and hospitalizations.

### Specimen collection

Participants were asked to collect and return 26 weekly nasal swab specimens beginning November through January. Prepackaged home collection kits were mailed to participants, and parents/guardians collected anterior nasal swabs (OMR-110, Genotek), placed swabs in a stabilizing solution and returned via pre-paid U.S. Postal Service envelopes. Upon receipt in the Thielen lab, specimens were aliquoted into cryovial tubes and stored at -80 [prior to analysis and sequenced in bulk.

### V4 16S rRNA Gene Amplification, Illumina Sequencing, and Microbiome Analysis

All samples were subjected to DNA extraction (DNeasy PowerSoil Pro HTP, Qiagen), targeted enrichment of the V4 region of the 16S rRNA gene V using the V4_515F_Nextera and V4_806R_Nextera primers [35], normalized with SequalPrep kits (Invitrogen) [36], pooled to create the sequencing library, and purified using AMPure XP magnetic beads (Beckman Coulter) and sequenced an Illumina MiSeq using a 600-cycle v3 sequencing kit (Illumina, San Diego, CA), yielding 300-bp paired-end sequence read by the University of Minnesota Genomics Center (UMGC) using established protocols.

16S rRNA sequences were processed using the QIIME2 pipeline from the gopher pipelines repository of the Minnesota Supercomputing Institute (MSI) (https://bitbucket.org/jgarbe/gopher-pipelines/wiki/qiime2-pipeline.rst). Briefly, reads were analyzed for quality using FastQC v. 0.11.9 and trimmed of primer and adapter sequences using cutadapt v. 0.40. Alignment to the bacterial genomes was performed using the SILVA 128 full-length reference database and QIIME 2 v. 2020.2. Post-alignment, read counts ranged from 43,784 to 282,885 per sample, with an average of 165,956 reads. Using the phyloseq and decontam packages in R, the dataset was filtered of contaminants with the prevalence method (threshold = 0.5), and then filtered for taxa with a prevalence across >10% of samples. Finally, the taxonomic data was agglomerated to the genus level, producing an OTU table that was composed of 65 taxa.

### Full 16S rRNA Gene Amplification, Oxford Nanopore Sequencing, and Microbiome Analysis

DNA was extracted from 200 µL of nasal swab in stabilization medium (OMR-110, DNAgenotek) using the ZymoBIOMICS DNA Miniprep Kit (Zymo Research) with minor modifications. Each extraction included water as a negative control and the ZymoBIOMICS Microbial Community Standard (D6300, Zymo Research) as a positive control. Cells were mechanically disrupted by vortexing for 40 minutes at maximum speed. DNA was eluted in 60 µL DNase/RNase-free water, quantified with the Quantus fluorometer (Promega), and stored at −80 °C. Full-length 16S rRNA genes were amplified using the 16S Barcoding Kit 24 (SQK-16S114.24, Oxford Nanopore Technologies) with primers 27F and 1492R. To accommodate low microbial biomass, 15 µL of DNA and 30 PCR cycles were used. PCR products were quantified, pooled to 50 ng per sample where possible (positive controls diluted 5×), and sequenced on a MinION Mk1B flowcell (FLO-MIN114) with high-accuracy basecalling (Min Q score = 9). Sequencing data was analyzed with the EPI2ME wf-16s workflow (v1.6.0, Oxford Nanopore Technologies), using the minimap2 classifier, min_read_qual = 12, max_len = 1800, min_len = 1200, min_percent_identity = 95, and min_ref_coverage = 90. The selected database_set was “ncbi_16s_18s”

### Viral Nucleic Acid Extraction, Reverse Transcription, Sequencing, and Identification

Total nucleic acid was extracted from respiratory swabs using the AllPrep PowerViral DNA/RNA Kit (Qiagen) without DNase treatment. An unused swab served as a negative extraction control, and NATtrol™ Respiratory Panel 2.1 (Zeptometrix) was included as a positive control. Reverse transcription, library preparation, and targeted enrichment were performed using the Illumina RNA Prep with Enrichment Tagmentation protocol with a target input of 50 ng total nucleic acid per sample. RNA was reverse transcribed using random hexamers, followed by second-strand synthesis, bead-linked transposome–mediated tagmentation, and PCR amplification with unique dual indices. Target enrichment was performed using the Illumina Respiratory Pathogen ID/AMR Oligo Panel with hybrid capture and post-capture PCR amplification. Final libraries were bead-purified, quantified, pooled at equimolar concentrations, and sequenced on an Illumina platform. Viral identification was performed using the Explify RPIP Data Analysis pipeline (BaseSpace) for all detected viral pathogens were included regardless of read count. Viruses were classified as acute (e.g., influenza, SARS-CoV-2, rhinovirus, parainfluenza, metapneumovirus, adenovirus, and seasonal coronaviruses) or chronic (e.g., CMV, HHV6, and EBV) based on established infection patterns.

### Alpha (measures of within sample diversity) and beta diversity (similarity or dissimilarity between two samples) analysis of microbiome samples

Microbial community composition was analyzed at the genus level using principal coordinates analysis (PCoA). Taxonomic abundances were CSS-transformed, and Bray–Curtis dissimilarities were computed to generate a sample-wise distance matrix. Ordination was performed using the ordinate() function from the phyloseq package with the ‘PCoA’ method. Shannon diversity and Chao1 richness of all samples was determined using the estimate_richness function from phyloseq.

### Pairwise associations of virus presence and microbiome community composition

Associations between viral presence and microbiome community structure (taxonomic PC1, taxonomic PC2, Shannon diversity) were evaluated using Wilcoxon rank-sum tests. For each donor (A-D), longitudinal samples were treated as replicates. Microbiome ordination values and Shannon diversity were tested separately against viral exposure variables, including overall viral presence, acute and chronic virus presence, and specific viral taxa. To ensure sample balance, tests were only performed when virus presence or absence represented at least 25% of the data. Wilcoxon rank-sum tests were applied to compare distributions of microbiome features between exposure groups, and resulting p-values were corrected for multiple testing using false discovery rate (FDR) adjustment.

### Differential abundance of taxa in response to virus presence

Prior to testing, longitudinal samples were aggregated by donor (A with B, and C with D) to increase statistical power. Differential abundance testing of taxa was performed using the MaAsLin2 framework. For each donor group, models were run separately with viral exposure variables as the primary fixed effect. These included overall viral presence, acute versus chronic infection status, and specific viral species. Scaled sequencing depth was included as a covariate, and donor identity (A-D) was modeled as a random effect. Taxonomic abundances were collapsed to the genus level, normalized using total sum scaling (TSS), and log-transformed before linear modeling. Associations were tested using linear models as implemented in MaAsLin2, and results were considered significant at a false discovery rate (FDR) of <10%.

## Results

### Cohort Design and Characteristics

Over three successive winter respiratory seasons, 64 participants were enrolled: 8 in 2021–2022, 22 in 2022–2023, and 34 in 2023–2024. Of these, 63 completed the baseline enrollment questionnaire, and 55 provided at least one survey (Figure 1). To evaluate the integration of novel diagnostic methods for concurrent microbiome and targeted virome analysis, we selected four participants from the 2021–2022 season for this pilot study. Key demographic characteristics of this subset are summarized in Table 1.

**Figure 1:**
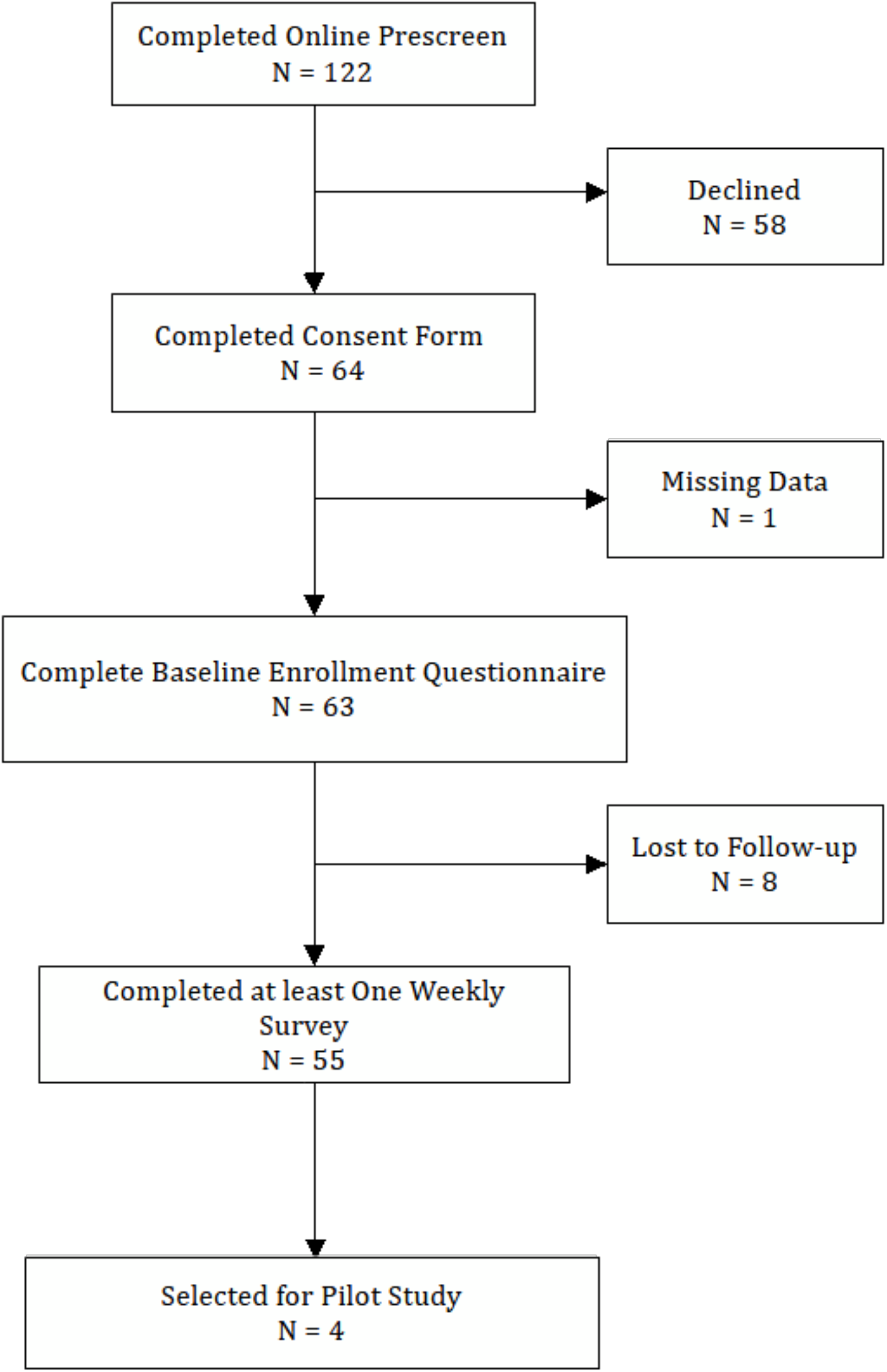
Participant enrollment and study flow: Flowchart depicting the stages of recruitment, eligibility screening, enrollment, and the longitudinal timeline of sample collection.

**Table 1:** Participant demographics: Demographic and clinical characteristics of the participants selected for pilot microbiome and virome analyses are summarized. Participants C and D are siblings living in the same household.

| Variables | A | B | C<sup>a</sup> | D<sup>a</sup> |
| --- | --- | --- | --- | --- |
| Age at Enrollment (5-month ranges)<br><sup>b</sup> | 21-25 | 21-25 | 41-45 | 1-5 |
| Sex | Female | Male | Male | Male |
| Self-identified Race | White, Non-Hispanic | White, Non-Hispanic | White, Non-Hispanic | White, Non-Hispanic |
| Number of Children in Home | 2 | 3 | 2 | 2 |
| Number of Adults in Home | 2 | 1 | 1 | 1 |
| Childcare Outside of Family | Yes | Yes | Yes | Yes |
| Mode of Delivery | Vaginal Delivery | Vaginal Delivery | Vaginal Delivery | Vaginal Delivery |
| Primary Nutrition (first three months of life) | All Breast Milk | All Breast Milk | Mostly Formula | Mostly Formula |
| Household Smoking Status | No | No | No | No |
| Pets | Yes | Yes | No | No |
| If Yes, Type | Dog | Reptile | -- | -- |

<sup>a</sup> Same household
<sup>b</sup> As of November 1<sup>st</sup> 2021

### Microbiome dynamics and viral detections

A total of 91 longitudinal specimens (15–26 per participant), including two positive and one negative control, underwent paired 16S rRNA (V4) sequencing and targeted-enrichment respiratory pathogen analysis for broad viral detection. As shown in Figure 2B, the nasal microbiomes of participants A and B were characterized by low diversity and the dominance of a few key taxa. These participants exhibited multiple rapid temporal shifts, reflecting high variability in microbial composition. *Moraxella* spp. were consistently abundant, with a median relative abundance of 27.66% (range: 6.91–70.31%) in participant A and 43.96% (range: 4.55–92.77%) in B. In participant A, *Haemophilus* spp. (median: 22.21%; range: 0.21–68.83%) and *Dolosigranulum pigrum* (median: 8.31%; range: 0.01–33.14%) exhibited an inverse temporal relationship. Similar patterns were observed in participant B, with median abundances of 1.42% (0.07–63.68%) and 14.74% (0–36.50%), respectively (Fig. 2B, Supplementary Fig. S1). Furthermore, several study weeks were characterized by transient spikes in *Streptococcus* spp. abundance. In contrast, siblings C and D possessed more diverse microbial communities characterized by a broader range of taxa. Their microbiomes exhibited greater week-to-week stability and fewer pronounced taxonomic spikes compared to participants A and B. *Streptococcus* was the predominating genus in both siblings (C: median 21.10%, range 12.73–32.82%; D: 27.13%, range 17.16–37.88%). Other shared dominant taxa included *Haemophilus* spp. (7.99% and 8.08%, respectively), *Gemella* spp. (7.24% and 6.10%), *Leptotrichia* spp. (3.85% and 6.90%), and *Neisseria* spp. (5.83% and 6.53%). Notably, *Veillonella* spp. and *Alloprevotella* spp. were considerably more abundant in sibling D (median 12.28% and 9.40%, respectively) than in sibling C (median 4.08% and 2.27%, respectively). Principal coordinate analysis (PCoA) of principal coordinate analyses (PCoA) using Bray–Curtis distances demonstrated strong separation between participants A/B and C/D, with the sibling pair C and D showing greater similarity in microbial composition than the unrelated participants A and B (Fig. 2C).

**Figure 2.**
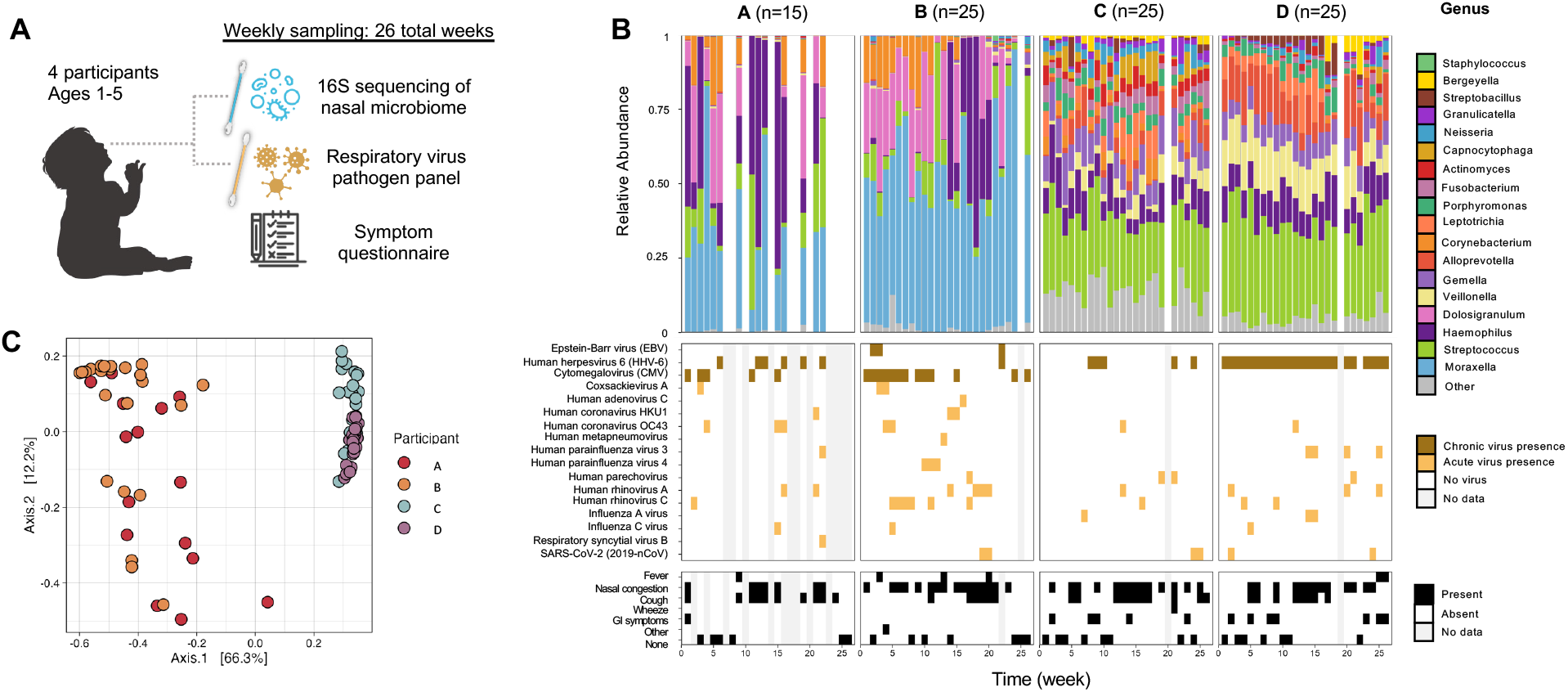
Study cohort overview and longitudinal microbiome-virome profile. (A) Schematic representation of the study cohort and sample collection timeline. (B) Top panel: Genus-level relative abundance of the bacterial microbiome. Samples are grouped by participant (A-D), with the total number of samples (n=) indicated above each section. White columns represent weeks where no samples were collected. The “Other” category includes taxa with a median relative abundance <0.05% (participants A/B) or <0.5% (participants C/D). Middle panel: Longitudinal viral detection. Chronic and acute viruses are indicated by brown and gold tiles, respectively; gray tiles denote weeks without sample collection. Bottom panel: Reported clinical symptoms. Black and white tiles indicate the presence or absence of symptoms, respectively; gray columns represent weeks with missing symptom data. (C) Principal Coordinates Analysis (PCoA) of genus-level beta diversity across all participant samples.

Viral analysis identified human herpesvirus 6 (HHV-6; 36 detections) and cytomegalovirus (CMV; 18 detections) as the most prevalent viruses, followed by human rhinovirus A and human rhinovirus C (HRV-A and HRV-C; 10 detections each) and human coronavirus OC43 (7 detections). Other detected pathogens included SARS-CoV-2 (n=6), human parainfluenza virus 3 (HPIV-3, n=5), human parechovirus (n=4), coxsackievirus A (n=3), and Epstein-Barr virus (EBV; n=3) (Fig. 2B). Chronic viral shedding was identified in 54 samples, while acute infections were detected in 42. Coinfections involving both chronic and acute viruses occurred in 26 samples. Furthermore, 17 samples contained multiple acute viruses, most frequently involving combinations of HRV-A (n=9), HCoV-OC43 (n=6), and HPIV-3 (n=5). No viruses were detected in 20 specimens.

Parental symptom reporting was frequent throughout the study period, with a total of 125 symptomatic reports across the four participants (A: n=17; B: n=32; C: n=39; and D: n=37). The most prevalent clinical manifestations were nasal congestion (n=48), cough (n=31), and gastrointestinal symptoms (n=13). Conversely, participants were asymptomatic for 25 collection dates.

### Associations between microbiome diversity and composition and virus detection

To quantify the major microbial taxa changing over time within individuals, we generated PcoA plots from Bray–Curtis distances for each participant (Fig. 3A). These ordinations captured within-individual variation, with dominant axes driven by genera such as *Haemophilus, Streptococcus, Dolosigranulum, Moraxella, Veillonella*, and *Alloprevotella*. We assessed genus-genus associations and positive correlations were observed between *Corynebacterium* and *Dolosigranulum, Capnocytophaga-Fusobacterium, Alloprevotella-Veillonella, Corynebacterium Helcococcus* (and *Bergeyella-Fusobacterium*. Additionally, we identified negative correlations between *Bergeyella-Veillonella, Corynebacterium Haemophilus*, and *Dolosigranulum-Haemophilus, Dolosigranulum-Moraxella* and *Fusobacterium-Veillonella*.

**Figure 3.**
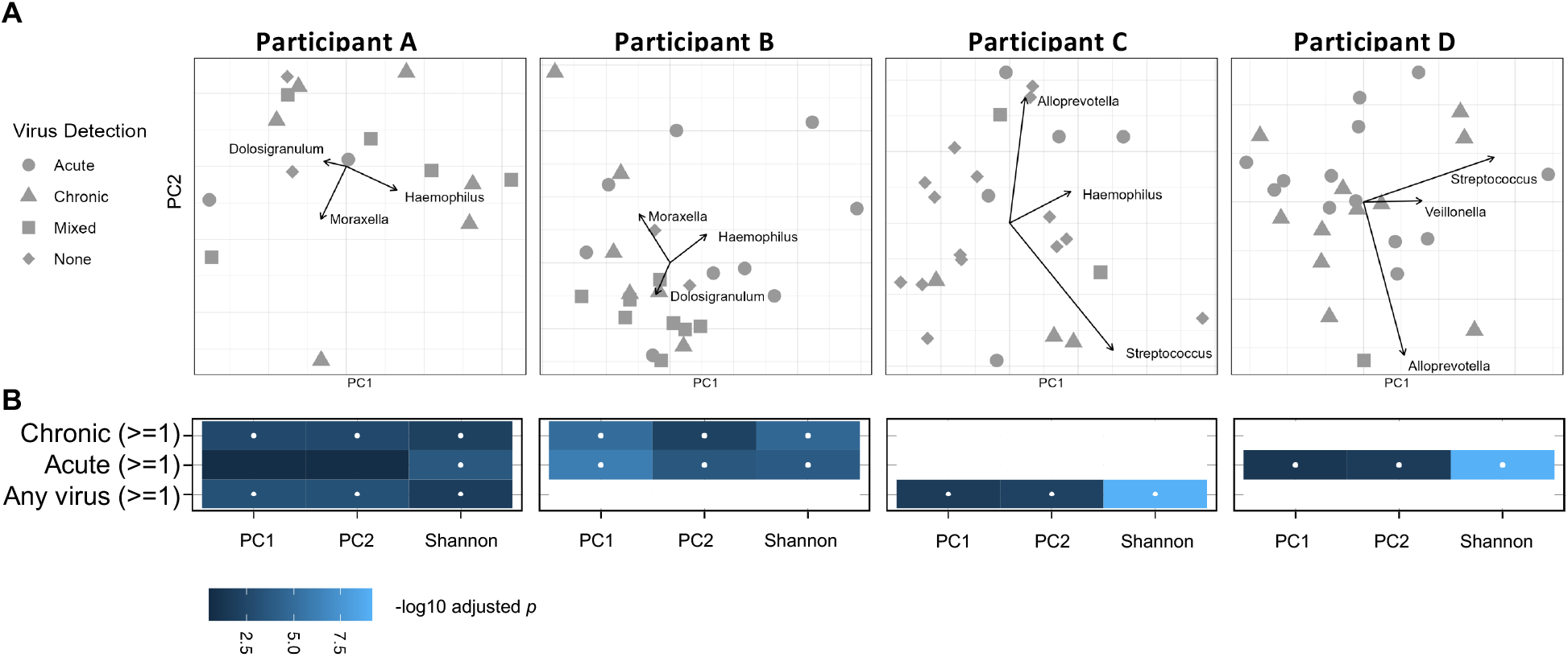
Drivers of microbiome variation and virus-microbiome correlations. (A) Principal Component Analysis (PCA) of genus-level microbial communities. Samples are color-coded by participant, with PC1 and PC2 representing the primary axes of variation. Red vectors indicate the top three taxa (eigenvectors) driving the observed clustering.(B) Heatmap showing correlations between viral summary features (y-axis) and microbiome diversity metrics (x-axis). Shading intensity represents the *p*-value, with significant associations (*p* < 0.05) denoted by a white dot. White tiles indicate feature pairs with insufficient or unequal distributions that were excluded from statistical testing.

**Figure 4.**
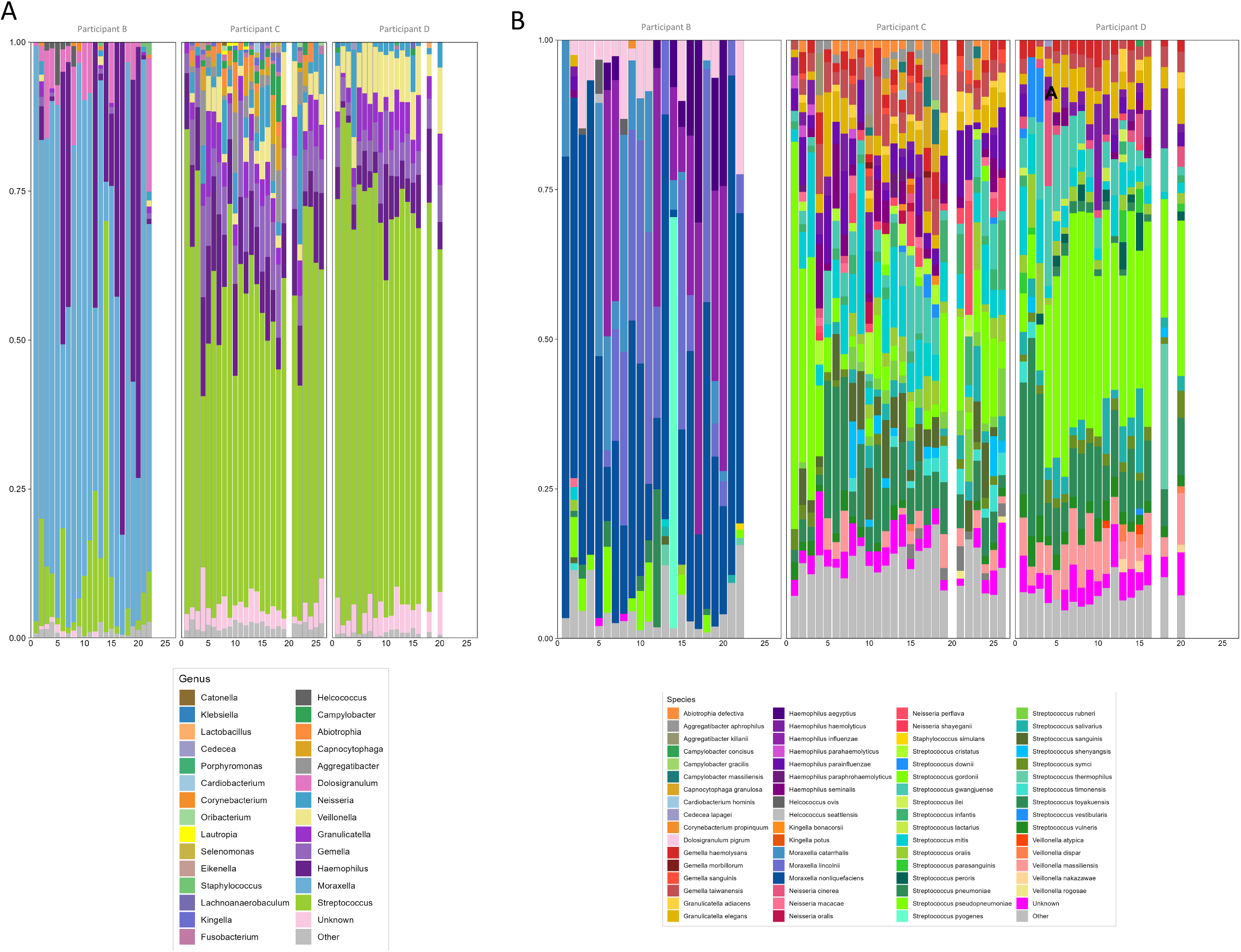
Longitudinal profile of full-length 16S rRNA-based microbial abundance. Relative abundance of bacterial taxa over time (x-axis) at the (A) genus level and (B) species level. White columns indicate weeks where no samples were collected. The “Other” category comprises taxa with a median relative abundance <0.05% for genus-level profiles and <0.01% for species-level profiles.

We next investigated the association between viral infections and microbial community structure by correlating viral status (binary presence/absence of acute or chronic infections) with longitudinal diversity metrics and taxonomic composition. Across all participants, α-diversity (Shannon index) fluctuated over time and was generally reduced during viral events. Baseline diversity differed markedly between individuals: A and B exhibited lower and more variable diversity (Shannon indices: 0.74–1.77 and 0.31–1.72, respectively), while siblings C and D maintained consistently higher diversity (2.25–2.98 and 1.73–2.54, respectively). Wilcoxon rank-sum tests indicated that virus-positive samples often exhibited significantly altered diversity, with the most pronounced differences occurring during acute viral detections (Fig. 3B). Linear mixed-effects modeling confirmed this trend: acute infections were associated with a significant reduction in Shannon diversity compared to infection-free weeks (β = −0.33, 95% CI: −0.58 to −0.08, p = 0.015). In contrast, neither chronic infections (β = −0.18, 95% CI: −0.44 to 0.07, p = 0.17) nor mixed acute/chronic infections (β = −0.21, 95% CI: −0.47 to 0.04, p = 0.12) significantly impacted diversity. A similar pattern was observed for species richness; Chao1 estimates were significantly reduced during acute infections (β = −33.2, 95% CI: −55.4 to −11.5, p = 0.004), whereas chronic (β = 3.3, 95% CI: −19.7 to 24.4, p = 0.77) and mixed infections (β = −6.9, 95% CI: −30.1 to 14.5, p = 0.54) showed no significant effects.

### Genus- and species-level resolution with full-length 16S rRNA sequencing using the Oxford Nanopore Technologies platform

A primary limitation of current short-read 16S rRNA sequencing protocols is the reliance on the V4 hypervariable subregion. This restriction often limits taxonomic resolution to the genus level, whereas meaningful clinical interpretation frequently necessitates identification at the species or strain level. To overcome these resolution constraints and leverage recent technological advances, we performed a comparative analysis using full-length 16S rRNA sequencing via Oxford Nanopore Technologies (ONT) alongside V4 sequencing of the same specimens. Of the original 91 samples, 65 retained sufficient material for full-length sequencing; participant A was excluded from this analysis due to sample depletion.

Consistent with the V4 findings, *Moraxella* spp. remained the most dominant genus in participant B (median 68.73%, range 6.56–96.58%), occurring in all samples. Full-length 16S rRNA analysis further identified *Streptococcus* spp. (median 6.30%, range 0.17–69.03%), *Dolosigranulum pigrum* (2.99%, 0–22.43%), and *Haemophilus* spp. (0.97%, 0.01– 82.64%) as the next most abundant taxa. A notable divergence from the V4 data was the ranking of *Dolosigranulum pigrum*, which appeared less dominant in the full-length analysis despite being the second most abundant genus in the V4 dataset. However, the inverse relationship between *Haemophilus* spp. and *Dolosigranulum pigrum* observed in the V4 data remained evident in the full-length results, though this trend warrants further validation in a larger cohort. At the species level, three distinct taxa contributed to the *Moraxella* population in participant B. *Moraxella nonliquefaciens* was the most frequently detected, followed by *M. lincolnii*. In contrast, *M. catarrhalis* reached high relative abundance (>10%) in only six non-consecutive weeks. Within the *Haemophilus* genus, *H. influenzae* and *H. aegyptius* predominated and were typically co-detected within the same study weeks. While members of the *Streptococcus* genus generally maintained a low combined relative abundance (median 6.3%; range 0.17–69.03%), we observed two significant episodic expansions: *Streptococcus pneumoniae* peaked at 23.1% during week 12, and *Streptococcus pyogenes* reached 68.7% in week 14. Finally, *Dolosigranulum pigrum* abundance remained variable, ranging from 0% to 22.4% (median 3.0%).

Consistent with the V4 data, *Streptococcus* was the predominant genus for both siblings C and D (median 51.91% and 67.21%; ranges 28.75–81.38% and 56.60–86.90%, respectively). Both siblings shared a core group of top genera— including *Haemophilus* spp., *Veillonella* spp., *Gemella* spp., *Granulicatella* spp., and *Neisseria* spp.—whereas *Dolosigranulum* was either absent or negligible. While these results largely mirrored the V4 profiles, *Alloprevotella* (which ranked third in the V4 analysis for participant D) was undetected by full-length 16S sequencing.

At the species level, the *Streptococcus* population was highly diverse. *S. pseudopneumoniae* was the most abundant, followed by *S. toyakuensis, S. mitis*, and *S. gwangjuense*; collectively, these taxa accounted for a median of 38.78% (range 6.75–70.57%) of the total community across both siblings. Notably, the major pathogen *S. pneumoniae* was rare, maintaining low relative abundances in both participant C (<0.08%) and D (<2.16%). The *Haemophilus* community was primarily composed of *H. parainfluenzae, H. haemolyticus*, and *H. seminalis*; in contrast to participant B, *H. influenzae* was absent in these siblings. Other species-level dominants included *Veillonella massiliensis, Granulicatella elegans*, and *Gemella taiwanensis* followed by *G. haemolysans*.

## Discussion

Longitudinal human cohort studies are essential for advancing our understanding of respiratory microbiome dynamics and viral epidemiology. No animal model currently recapitulates these interactions; most animal models lack susceptibility to the full spectrum of human respiratory pathogens, harbor distinct indigenous microbiota, and manifest divergent clinical outcomes in response to infection [37]. Although prior studies have characterized the pediatric upper respiratory microbiome and its associations with specific pathogens, only one longitudinal study has examined microbiome–virus correlations in infants under one year of age [15], leaving the dynamics in children aged one to five years largely unexplored. This represents a critical gap in understanding how respiratory viruses shape microbial communities across early childhood.

To address this critical knowledge gap, we conducted a three-year longitudinal study across successive winter seasons, collecting symptom surveys and nasal swabs from 55 participants under five years of age. For this pilot analysis, we utilized a subset of 91 longitudinal specimens from four participants (2021–2022 season) to integrate standard V4 16S rRNA sequencing with full-length 16S rRNA analysis and broad viral detection via target-enrichment panels.

Importantly, this study demonstrates the feasibility of longitudinal, home-based specimen collection in children under five—a population historically underrepresented in clinical research [38]. By utilizing child-friendly protocols and targeted parental engagement, we successfully collected high-quality nasal swabs remotely without the requirement for immediate refrigeration. Despite being parent-collected and transported via standard mail, these specimens yielded robust, high-fidelity sequencing data. The ability to perform at-home collection and ambient-temperature transport distinguishes this study from prior large-scale cohorts [39] and facilitates broader, geographically diverse enrollment by eliminating the logistical burden of in-person clinical visits.

At the genus level, distinct ecological profiles emerged: two participants were dominated by *Moraxella* spp., *Haemophilus* spp., and *Dolosigranulum pigrum*, while the siblings were enriched in *Streptococcus* and, to a lesser extent, *Haemophilus, Alloprevotella* spp., *Veillonella* spp., *Gemella* spp., and *<u>Leptotrichia</u>* spp.. These patterns are largely consistent with previous longitudinal data, though our detection of *Alloprevotella* spp., *Veillonella* spp., *Gemella* spp., and *Leptotrichia* spp. is particularly noteworthy. These genera are more commonly associated with the oral microbiome and have been reported to be enriched in younger children, those delivered via caesarean section, or breastfed infants. Furthermore, this study recapitulated previously described inter-genus associations, including the frequent co-occurrence of *Corynebacterium* spp. and *Dolosigranulum pigrum. D. pigrum* remains of significant clinical interest, as it has been shown to inhibit colonization by major pathogens such as *Streptococcus pneumoniae* and *Staphylococcus aureus* in vitro[40,41], highlighting its potential as a beneficial or therapeutic commensal in the pediatric airway [42].

A notable strength of this study is the demonstrated utility of long-read sequencing for high-resolution characterization of the nasal microbiome. At the genus level, V4 Illumina and full-length 16S rRNA Oxford Nanopore Technologies (ONT) approaches exhibited broad concordance in identifying dominant taxa; *Moraxella* spp. (participants A/B) and *Streptococcus* spp. (siblings C/D) consistently ranked as the most abundant genera across both platforms. However, despite this overall agreement on core community members, we observed variations in relative abundance estimates and taxonomic rank ordering. These discrepancies likely reflect methodological differences, including primer bias, sequencing chemistry, and classification-dependent effects. A detailed comparison of these platform-specific differences is provided in Supplementary Text S1.

Another key finding of our study is that full-length 16S rRNA sequencing provided species-level resolution unattainable through short-read V4 profiling, revealing critical distinctions between commensal and pathogenic taxa within the same genera. In participant B, for example, *Moraxella* populations were dominated by the typically commensal species *M. nonliquefaciens* and *M. lincolnii*. In contrast, the respiratory pathogen *M. catarrhalis* appeared only intermittently, reaching elevated abundances for a limited duration—a pattern suggesting transient colonization rather than stable dominance. Within the *Haemophilus* genus, the frequent co-occurrence of *H. influenzae* and *H. aegyptius* in participant B contrasted sharply with the predominance of opportunistic or non-pathogenic species (*H. parainfluenzae, H. haemolyticus*, and *H. seminalis*) in siblings C and D. These participant-specific profiles suggest varying levels of potential pathogenic burden. Notably, *Dolosigranulum* was entirely absent in the siblings who maintained a stable presence of non-pathogenic *Haemophilus*, potentially offering new insights into the known inverse relationship between these genera. Species-level resolution further uncovered episodic blooms of clinically significant pathogens, including *Streptococcus pneumoniae* and *S. pyogenes*, which would have been obscured had *Streptococcus* been analyzed solely at the genus level. Together, these results demonstrate that full-length 16S sequencing provides essential clinical value by resolving functionally distinct species, enabling a more precise interpretation of microbial dynamics and disease risk. Consequently, we intend to implement full-length ONT sequencing for all remaining samples in this cohort.

Consistent with prior household studies [29], we observed a high burden of acute respiratory viruses in our young participants, with rhinovirus and coronavirus being the most common. Importantly, the use of a novel target-enrichment approach enabled detection of persistent or recurrent shedding of herpesviruses—EBV, CMV, and HHV-6—which are not typically included in standard respiratory viral panels, as well as parvovirus B19, which has previously been reported as common [43,44]. Studies in adults with pneumonia have shown that herpesvirus detection is associated with differences in respiratory microbiome composition [45], highlighting a promising avenue for future investigation in pediatric populations. Acute viral infections—most commonly human rhinovirus and coronaviruses—coincided with transient reductions in Shannon diversity and shifts in community composition, whereas chronic viruses such as CMV and HHV-6 were associated with more moderate but persistent microbiome alterations.

While these findings provide valuable insights into pediatric respiratory microbiome–virome interactions, several limitations must be considered. First, this pilot analysis included only four participants, which limits the generalizability of the findings and the overall statistical power. Second, while parental symptom reporting was frequent, data were captured as binary metrics (presence/absence), which constrains the assessment of clinical severity or symptomatic burden. Furthermore, viral analyses relied on presence/absence data rather than quantitative viral load measurements. Because the hybrid-capture sequencing recovered only short genomic fragments, we were limited in our ability to distinguish between prolonged viral persistence and novel re-exposures. Finally, standard short-read 16S rRNA sequencing restricted taxonomic resolution to the genus level, masking the diversity of clinically significant species within complex genera such as *Haemophilus* spp., *Streptococcus* spp., and *Moraxella* spp..

Despite these limitations, this pilot study demonstrates the feasibility of high-frequency, longitudinal profiling of the nasal microbiome and virome in young children using fully remote collection and multi-omic next-generation sequencing. Our findings highlight the superior utility of full-length 16S rRNA sequencing in achieving species-level resolution, which is essential for characterizing the clinically significant interactions between specific commensals and respiratory viruses. Future investigations utilizing larger, genetically diverse cohorts, quantitative viral load measurements, and expanded high-resolution sequencing will provide definitive insights into the mechanisms linking viral infection, microbial community dynamics, and pediatric respiratory health. Ultimately, this approach establishes a robust foundation for mechanistic research and offers a scalable framework for integrated microbiome–virome surveillance in early childhood.

## Data Availability

All data produced in the present study are available upon reasonable request to the authors and will also be available online in the next version of the manuscript.

## Legends

**Table 1: Participant demographics**: Demographic and clinical characteristics of the participants selected for pilot microbiome and virome analyses are summarized. Participants C and D are siblings living in the same household.

## Acknowledgments

We gratefully acknowledge the study participants and their families for their time, commitment, and partnership in this research. We acknowledge the University of Minnesota Genomics Center for performing sequencing assays under a fee-for-service contractual agreement. We thank past and present members of the laboratory--Anwen Eslinger, Madeline Addington, and Idil Abdi--for their technical assistance, logistical support, and helpful discussions throughout the course of this study and University of Minnesota Pediatric Clinical Research Services staff Amy Kodet, Joanna Ly, Emily Graupman, and Melina Lee for clinical research support of the study.

