## Supplementary text 1 for "Longitudinal Weekly Surveillance of Respiratory Viruses and the Nasal Microbiome in Children Under Five (MINNE-LOVE Study)"

Supplementary text S1 – Comparing V4 Illumina and full-length ONT 16s sequencing results.

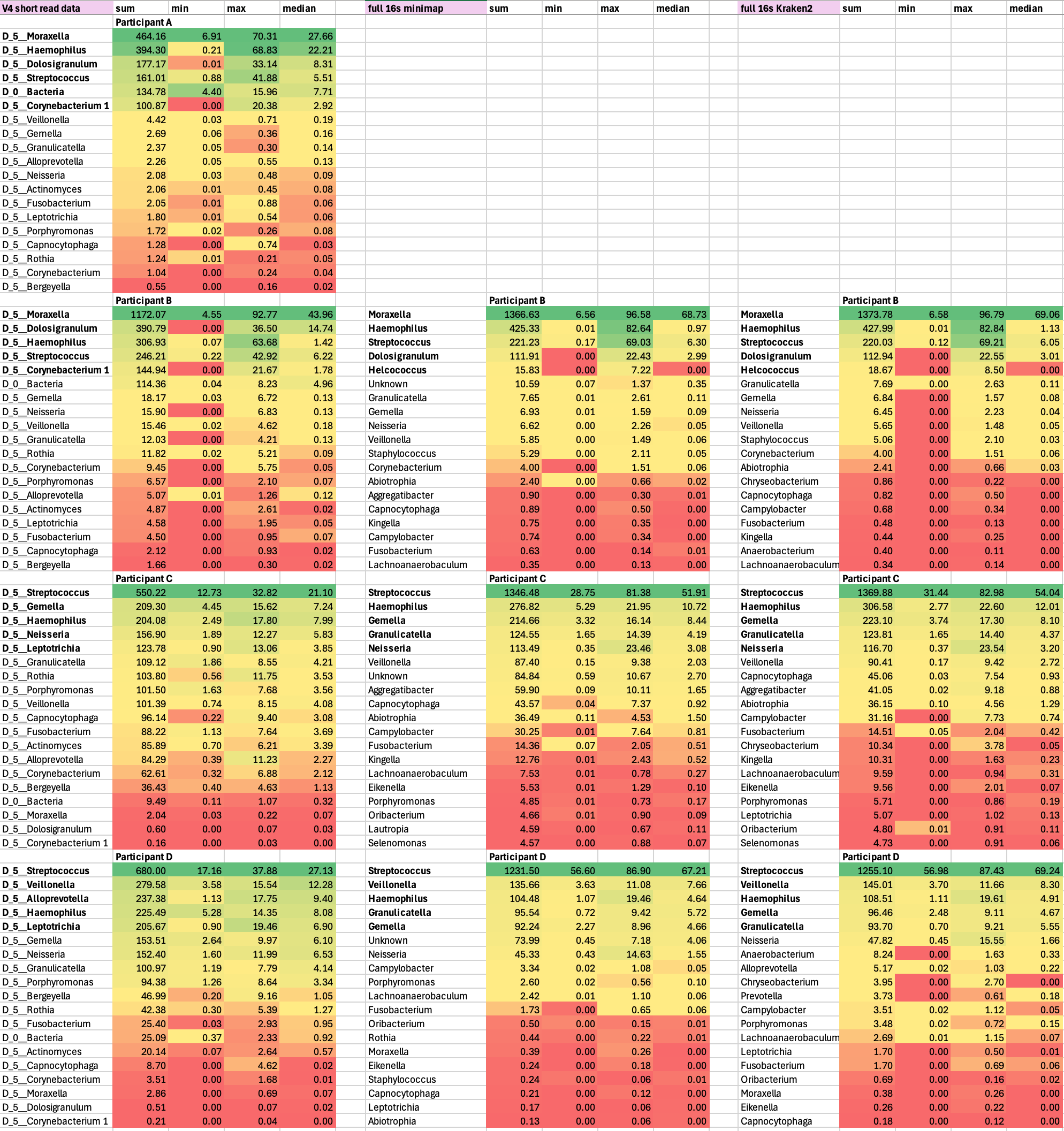

Figure S1. Comparison of V4 Illumina relative abundance data with full-length 16s (ONT sequence data for top 19 identified genera for each of the study participants, listing cumulative relative abundances, the relative abundance range over all sampled weeks, and the median relative abundance. No samples for participant A were available at the time of full-length 16s experiments.

Here we discuss some of the discrepant findings between V4 Illumina 16s sequencing and full-length ONT 16s sequencing for specific taxa. The full-length 16s data was analyzed with the minimap2 classifier (alignment-based) option in the EPI2ME wf-16s workflow (Oxford Nanopore Technologies). In addition, we also ran the analysis with the Kraken2 (k-mer based) classification to further consider method-based discrepancies with the V4 Illumina approach. For both the minimap2 and kraken2 approach the ncbi_16s_18s reference database was used. SILVA, the database used for V4 Illumina sequence analysis, is not suitable for species-level classification as it emphasizes conserved rRNA regions and contains a large proportion of entries annotated only to genus or higher taxonomic ranks. As a result, species-level assignments using SILVA often remain ambiguous or collapse closely related taxa into shared lineages, particularly for genera with high 16S sequence similarity such as Streptococcus, Haemophilus, and Neisseria.

Higher relative abundances for top genera *Moraxella* (participant B) and *Streptococcus* ( siblings C and D) with full-16s sequencing

Full-length 16S (approx. 1,500 bp) provides significantly higher resolution than the V4 region (approx. 250 bp). Short V4 reads often fail to distinguish between closely related species within *Streptococcus* and *Moraxella*, which can lead to "under-counting" if reads are discarded as ambiguous or misclassified at the genus level (PMID 36321824, 37623922). Database choice may additionally account for these observed differences as higher sequence matching success rates enabled by the NCBI database's extensive sub-genus taxonomic depth and species-level resolution reduces the proportion of sequences that remain unclassified compared to the more conservatively curated, genus-focused SILVA database (PMID: 34748542).

Lower *Dolosigranulum* relative abundance with full-16s sequencing (participant B)

*Dolosigranulum* was one of the abundant genera in participant A (0.01-33.14, 8.31) and B (0-36.50, 14.74) with V4 Illumina data. The substantially lower relative abundance of *Dolosigranulum* for participant B (0 – 22.43 %, median = 2.99 %) observed with full-length ONT 16S sequencing in conjunction with minimap2 likely reflects amplicon region and primer bias rather than biological absence. For example, studies comparing full-length and short-read protocols highlighted that the ONT forward primer 27F has lower binding efficiency for certain Firmicutes, including *Dolosigranulum* when compared to more robustly 515F/806R primers used for the V4 region (PMID: 37564874, 38934545). The kraken2-based analysis resulted in a similar relative abundance range (0.00-22.55 %, median = 3.01%).

Abundant *Alloprevotella* presence in participant D with V4 Illumina sequencing, but absent in full-16s sequencing (Minimap2 approach)

One of the top taxa in participant D samples with V4 Illumina, *Alloprevotella* (1.13-17.75, 9.40), was completely absent in the full-length 16s sequence data when the Minimap2 approach was used. While more research is needed to explain this observed difference, this likely results from a combination of primer binding bias, PCR efficiency differences leading to preferential amplification with the V4 primers, in addition to database differences. Interestingly, when reanalyzing the full-length 16s sequence data with the Kraken2 approach, Alloprevotella was detected (0.02-1.03 %, median = 0.22%), though less abundant than with V4 sequencing. Closely related *Prevotella* was also observed with the k-mer based kraken2 approach.

While the most abundant taxa were largely recovered across sequencing approaches, notable differences in taxon presence/absence and relative abundance were observed between short-read V4 Illumina sequencing and full-length 16S Oxford Nanopore sequencing. These discrepancies are consistent with known methodological effects rather than true biological variation. Primer selection and targeted hypervariable region strongly influence amplification efficiency and taxonomic resolution, while sequencing platform–specific error profiles, taxonomic classification algorithms, pipeline parameterization, and reference database choice further shape observed community composition. Previous benchmarking studies have demonstrated that short-read, region-trained classifiers and long-read, full-length alignment-based approaches can yield systematically different abundance estimates for the same taxa, underscoring the importance of methodological context when interpreting microbiome profiles.
